# Using the Public Health and Social Measures Severity Index for analysing government responses to COVID-19 in the WHO European Region

**DOI:** 10.1101/2023.11.21.23298818

**Authors:** Christoph Wippel, Lisa Owen, Dominic Cocciolone, Jussi Sane, Jennifer Addo, Christian Gapp, Aimee Latta, Sandra Lindmark, Tanja Schmidt

## Abstract

Public health and social measures (PHSM), such as adaptions to the operation of schools, businesses and workplaces, international travel measures and restrictions on gatherings and people’s movements, as well as individual preventive measures such as physical distancing, have been a vital set of tools utilized by many countries to mitigate the spread of SARS-CoV-2 virus throughout the COVID-19 pandemic. In January 2020, the WHO Regional Office for Europe started to systematically monitor, collect and categorize data on response measures taken by its Member States. In order to visualize and analyse the collected data, WHO developed a methodology for quantifying the response measures into an index, capturing the severity of these policy measures in terms of six key indicators. The aim of this article is to describe the methodology underlying the index, including data collection, categorization and calculation, in order to provide researchers and policy-makers with a better understanding of its application. Furthermore, it provides an overview and examples of possible applications of the index, as well as serve as a reference for subsequent research on the effectiveness of PHSM, including empirical studies and models that can help to guide health policies, their timing and severity.

## Introduction

On 30 January 2020 WHO declared the SARS-CoV-2 virus outbreak a Public Health Emergency of International Concern *(1)*. The subsequent characterization of the outbreak as a pandemic on 11 March 2020 *(2)* galvanized most governments around the world to make drastic policy decisions, introducing a wide range of, at times, highly restrictive measures, with the hope of stopping or slowing the transmission of the SARS-CoV-2 virus. These ranged from social measures, such as international travel restrictions, closure of schools, businesses, workplaces and restricting people’s movements and their ability to gather, to public health and infection prevention and control measures, such as testing, contact tracing and isolating cases. The simultaneous roll-out of a bundle of measures, collectively described as public health and social measures (PHSM), allowed most countries to slow the spread of the initial wave of the disease to some degree *(3)*. Throughout the pandemic, PHSM have remained a vital set of tools for governments and populations in reducing the spread of the virus, even with the development and distribution of effective vaccinations against COVID-19 *(4)*. While most countries’ governments have implemented a combination of different PHSM in response to COVID-19, the timing (initial implementation and length of measures), degree, and scope of individual and societal PHSM responses have varied significantly *(5,6)*.

WHO has advised that PHSM should be implemented and adjusted according to local settings and conditions with the intent to curb transmission while keeping the negative impact on people’s social, mental, and physical well-being, as well as on the economy, at a minimum *(5)*. These decisions should be made based on evidence derived from methodologically rigorous research using context-specific yet comparable data *(7)*. To strengthen the evidence base for decision-making, it is crucial to have standardized systems in place that monitor and track governments’ response measures and enable the comprehensive collection and the meaningful analysis of data. A number of regional and global initiatives have attempted to address this need during the COVID-19 pandemic *(8–10)*.

In January 2020, the WHO Regional Office for Europe’s COVID-19 Incident Management Support Team (IMST) began monitoring response measures implemented by its member states, making it one of the first initiatives to systematically collect data on PHSM. With over 30 000 entries, encompassing the policy responses of 53 Member States, the PHSM database comprises one of the most comprehensive publicly available collections of PHSM data *(11)*. In order to quantify, visualize, and analyse the collected data, the Regional Office developed the PHSM Severity Index (SI), which is a composite score capturing a time series of severity of policy measures in six key areas of PHSM.

The SI has been continually used by the IMST throughout the pandemic to provide country support and guidance. In conjunction with community transmission indicators, the SI has also informed the situational assessment of national PHSM adjustment efforts with the help of the PHSM Calibration Tool *(12)*. Within this tool, the SI is displayed alongside case incidence, mortality rate and vaccination uptake, so that policy-makers who utilize the online tool can compare these indicators in order to facilitate decision-making *(13,14)*.

PHSM data was first made publicly available in the *COVID-19 Situation in the WHO European Region* dashboard in November 2020 *(15,16)*. The first iteration of the methodology for the SI was published in December 2020 *(17)* and has been continually updated as the pandemic evolved *(8)*.

The aim of this article is to describe the methodology underlying the index, including its data collection, categorization and calculation, in order to provide researchers and policy-makers with a better understanding of its application. Furthermore, it provides an overview and examples of possible applications of the index, as well as serving as a reference for subsequent research on the effectiveness of PHSM, including empirical studies and models that can help to guide health policies, their timing and severity.

## Methods

### Data collection

Countries were monitored daily, and data on PHSM were collected throughout the pandemic by a monitoring team consisting of five to seven members, starting in January 2020. Each team member was responsible for a group of countries, covering the 53 Member States of the WHO European Region. For each country, data on PHSM were systematically captured from four main sources: (i) government websites; (ii) WHO country office and direct member state reporting to WHO; (iii) international and national media sources; and (iv) thematic webpages on PHSM. PHSM data were stored in the PHSM measures database, categorised according to the seven categories and 42 subcategories of the WHO PHSM taxonomy and glossary *(11)*. The order of sources served as a hierarchical order of preference for data collection; i.e. if information on a specific PHSM was available from a government source, data was entered from that source, otherwise, the next source in the order was used, and so on. If conflicting information regarding timing or the nature of specific measures was identified from different sources, accuracy was assumed in the same order of preference, and data entered in the database accordingly. Each entry in the database can be regarded as a change in a country’s PHSM policy in a certain category; i.e. an entry was made if a measure was introduced, and a new entry was made if the policy of said measure was changed or lifted. In addition to categorising entries according to the taxonomy, the date of introduction, a short description of the measure, information on whether a measure was first introduced, reintroduced, extended, strengthened or lifted, and whether it applied to the whole country or on a subnational level were all recorded.

### Ordinal measures policy scale and binary scope scale

To allow for comparison, visualization and analysis of the qualitative data collected in the PHSM database, a method was devised to quantify the data in a standardized SI based on the severity of government measures for each day of the COVID-19 response. Six key categories were selected to comprise the index, including policies on masks, schools, businesses, gatherings, domestic movement and international travel. These six indicators were chosen because they represent common categories of large-scale measures implemented by a considerable number of countries throughout the WHO European Region throughout the pandemic. By considering a select number of common measures categories of the COVID-19 response, the SI allows for a standardized cross-country comparison. Each country was scored on an ordinal measures policy scale (henceforth referred to as “coding”) corresponding with the degree of severity of the response policy in each of the indicator categories. This ranged from no measures in place in a given category (0), to the most severe form of measures in that category (maximum ordinal scale value) for each day on a timeline based on the policy changes collected in the PHSM database (Tables Table 1). The severity score was applied to subsequent days until a change in policy occurred that caused a further change in the ordinal scale. An additional binary scope variable was recorded for each day and indicator, distinguishing whether the recorded severity was applicable to the whole country or population (general – 1) or only in a specific subnational locality or within a subpopulation (targeted – 0).⍰If measures differed within a country, the highest ordinal scale value of the targeted policy change was recorded. The indicators, as well as their ordinal scales were refined as the pandemic evolved and new information became available.

**Table 1.**
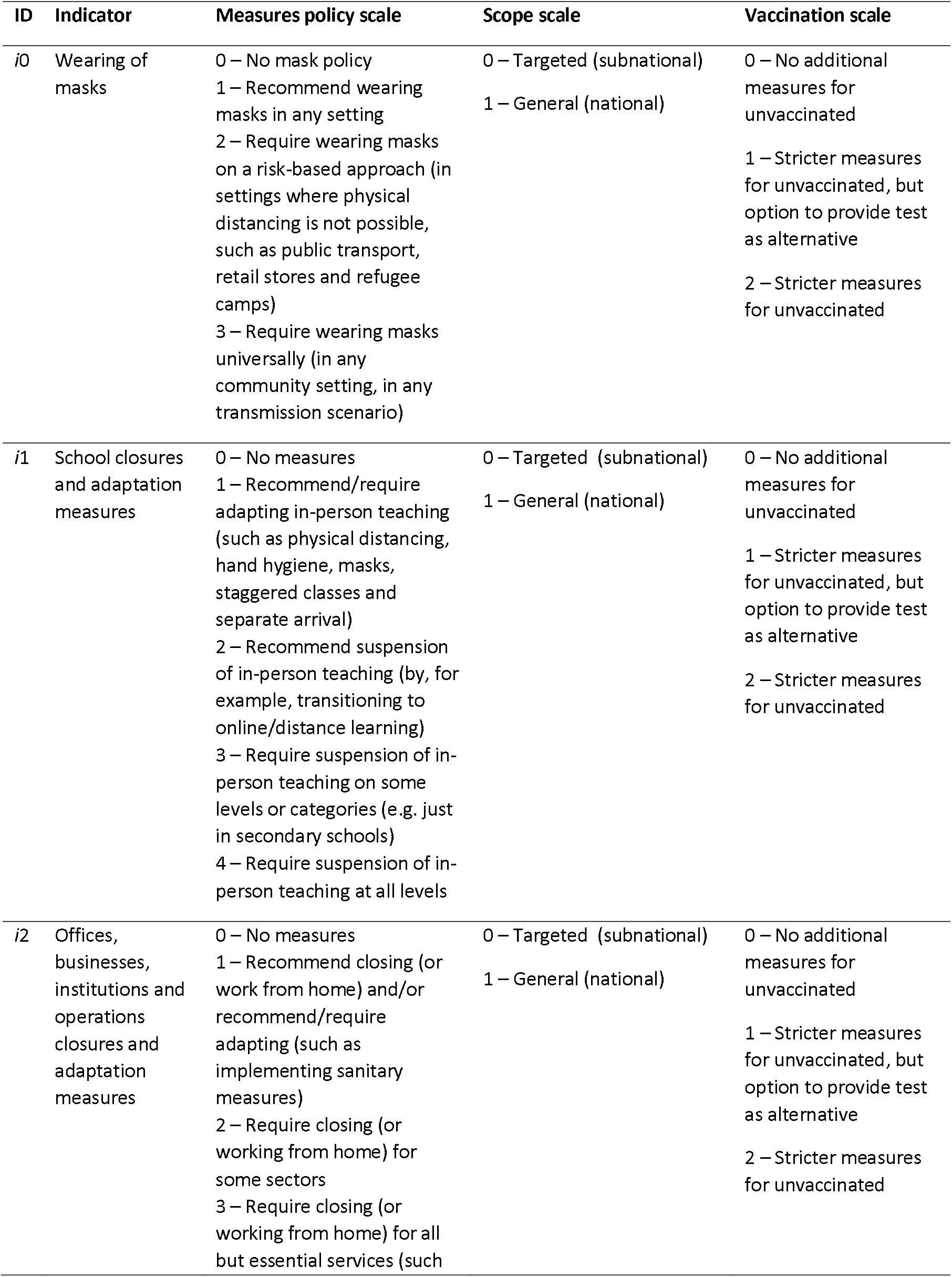
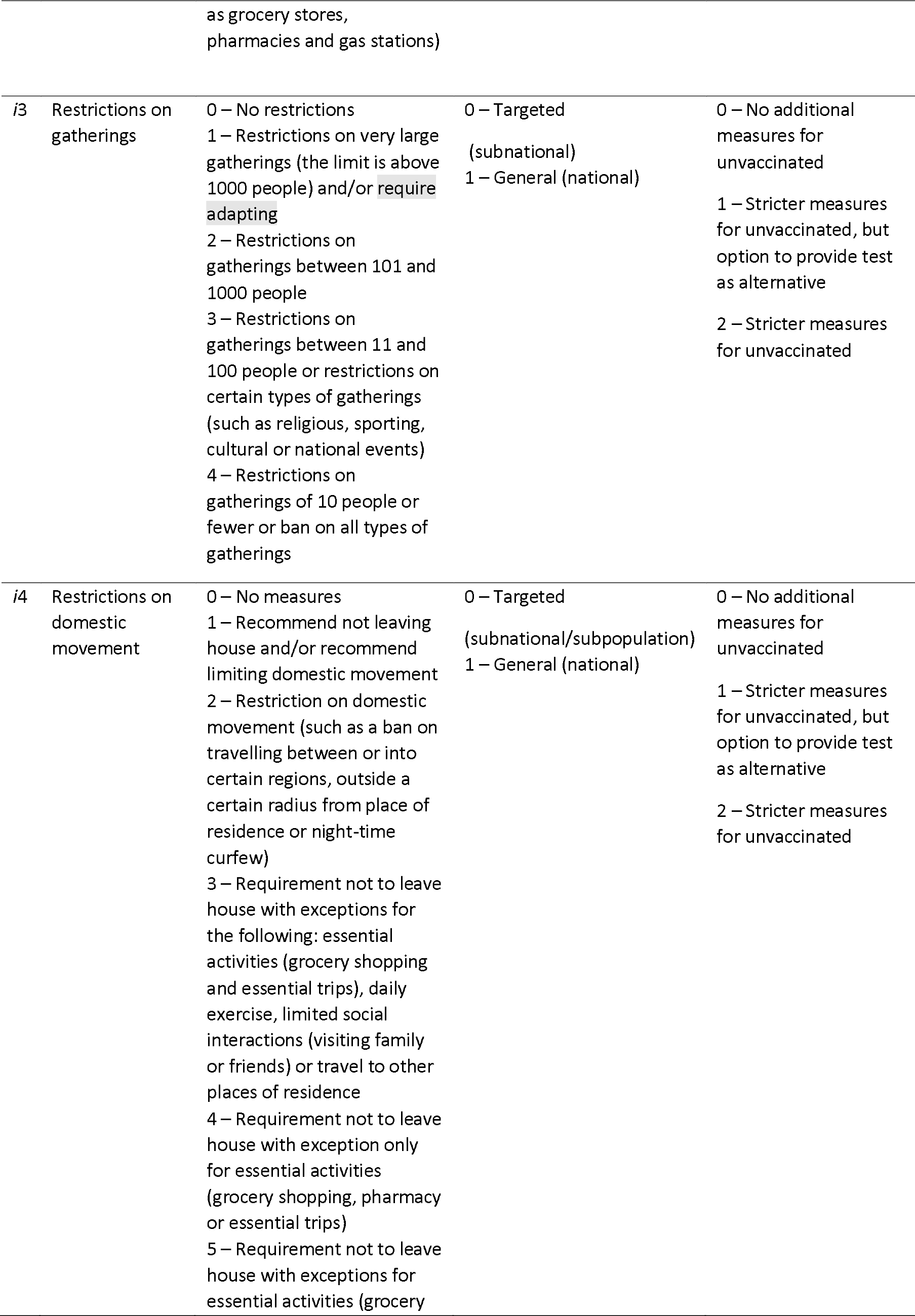
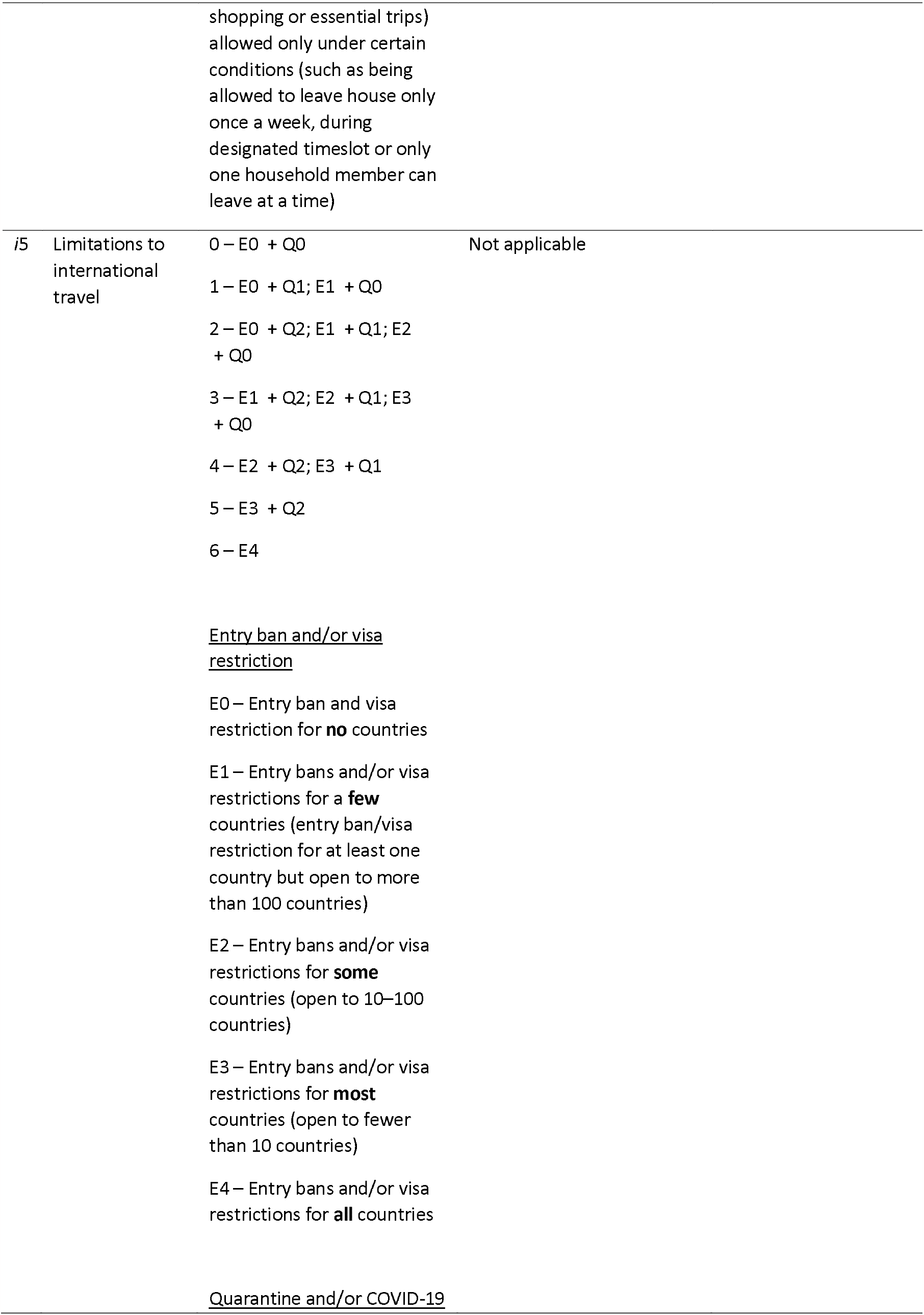
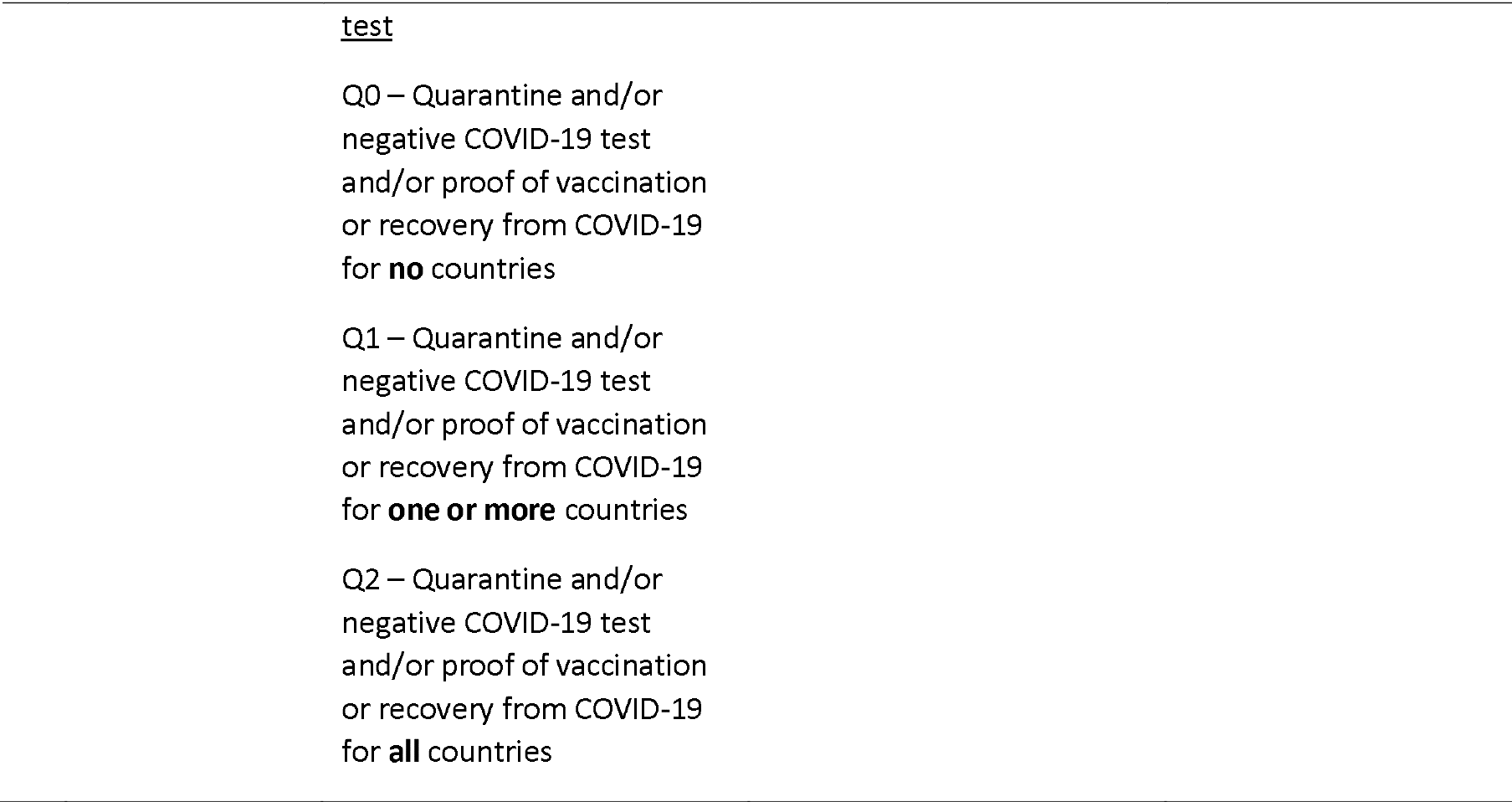
Ordinal and scope scales of the six PHSM indicators

### Ordinal vaccination scale

As the COVID-19 pandemic evolved, countries integrated COVID-19 vaccination into other PHSM policies and it became necessary to add an additional ordinal variable to the SI methodology to accurately reflect PHSM severity. The vaccination scale considers modifications of PHSM through vaccination policies; i.e. if more severe measures were implemented for unvaccinated people such as a lower limit on gatherings for those who could not show proof of having received a vaccination or having recently recovered from COVID-19, the vaccination scale increased the overall severity of the respective indicator. A score of 1 on the vaccination scale denotes more severe measures for the unvaccinated where the possibility existed of providing a recent negative COVID-19 test result as a way to avoid those measures, and a score of 2 if this possibility was not available.

### Coding principles

A set of general coding principles and specific principles for each indicator were defined to ensure consistency in coding across all countries and to avoid discrepancies within the SI, as outlined in the SI methodology published by the WHO Regional Office for Europe *(8)*. These principles detail which measures were coded, how they were coded and how the various measures contributed to severity scores. Providing these coding principles clarifies how government policies are translated into severity scores, and fosters a better understanding of the interpretation and application of the SI.

### Calculation of the index

For each indicator, an index was calculated taking into account the ordinal measures policy scale value, the binary geographic scope variable and the ordinal vaccination scale value for each day of the response. The overall SI was then calculated using the arithmetic mean of the indicator SIs for each day. The indicator score was calculated according to the following equation, where *n*_*i,d*_ is the value of the indicator score for the indicator *i* on any given day d. *m*_*i,d*_ represents the ordinal measures policy scale value for indicator *i* on any given day d. *S*_*i*_ is the possible scope for indicator *i* (1 for indicators *i0–i4* and 0 for indicator *i5*) and *s*_*i,d*_ is the binary value of the scope (0 or 1) for indicator *i* on any given day d. *M*_*i*_ represents the maximum value of the measures policy scale of indicator *i*.

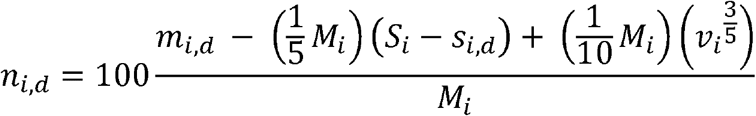

*n*_*i,d*_ = i<u>n</u>dicator score for a given indicator at a given day (0–100)

*m*_*i,d*_ = ordinal <u>m</u>easures policy scale value for a given indicator at a given day (e.g. 0, 1, 2, 3, 4…)

*M*_*i*_ = <u>M</u>aximum value of ordinal measures policy scale (if scale 0–5, then 5)

*S*_*i*_ = possible <u>S</u>cope value (1 if indicator includes scope, 0 if no scope)

*s*_*i,d*_ = <u>s</u>cope value for a given indicator at a given day (0 = targeted/regional, 1 = general)

*v*_*I, d*_ = ordinal <u>v</u>accination scale value for a given indicator at a given day (2 = stricter measures for unvaccinated, 1 = stricter measures for unvaccinated, but testing possible, 0 = no additional measures for unvaccinated)

If an indicator has a binary scope variable (that is, if a measure is targeted to a specific geographical region or group of persons), the calculation of the indicator score takes this into account by reducing the coded measures policy scale value by one fifth of the maximum value of the measures policy scale. If additional or stricter measures were in place in one category for people who were unvaccinated, the vaccination scale (*v*_*i*_) was added, accounting for the additional severity of the measure in the calculation of the index. The weighted reduction relative to the maximum scale ensures that the scope was weighted equally regardless of the number of levels in the measures policy scale of an indicator. The result is a normalized severity score between 0 and 100, with larger values representing more severe measures. Based on this formula, a measure that targets a specific region or group of people is less severe than an equally intense national-level measure that targets the entire population. The composite SI is the arithmetic mean value of the severity scores of all six indicators for any given day.

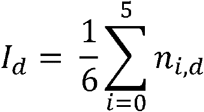

I^d^ = severity index score for any given day

### Validation

The methodology of the SI was validated in a process of iterative improvements based on continuous discussions with expert groups on different aspects of PHSM. Furthermore, validation exercises of the index were conducted to ensure the consistency and accuracy of coding whenever noteworthy changes were made to the measures policy scale and principles. As part of the validation exercises, a sample of countries was randomly assigned to independent validators who completed country coding based on the measures database without reference to the initial coding. The results of the recoding were then compared with the original coding to assess for congruence, and any deviations were discussed and addressed. When necessary, principles were added or refined to ensure that ambiguous and nonstandard measures were coded consistently in a similar manner across all countries.

## Discussion

The SI allows for in-depth analysis of country PHSM responses, cross-country comparisons through timelines and maps, and analysis of the relationship between PHSM and the epidemiological situation. These forms of analyses can address public policy questions related to PHSM responses, support policy-makers within countries and inform the development of specific policy-making guidance for current and future pandemic responses. Furthermore, the SI aims to provide insights into individual country PHSM responses, and how various key measures contribute to the comprehensive response. Specifically, visualizing national responses can provide clarity on the PHSM response by displaying the sequence and timing of PHSM implementation and the relative severity of the various measures *(15)*. Common PHSM strategies, outliers and emergent patterns can be identified through regional-level analysis and, when supported by country-specific examples, provide a comprehensive overview of the past and current regional situation. This allows countries to learn from various approaches to implementing PHSM and informs policy-making by enabling countries to better understand how their PHSM approach compares to other countries (Fig. 1).

**Figure 1.**
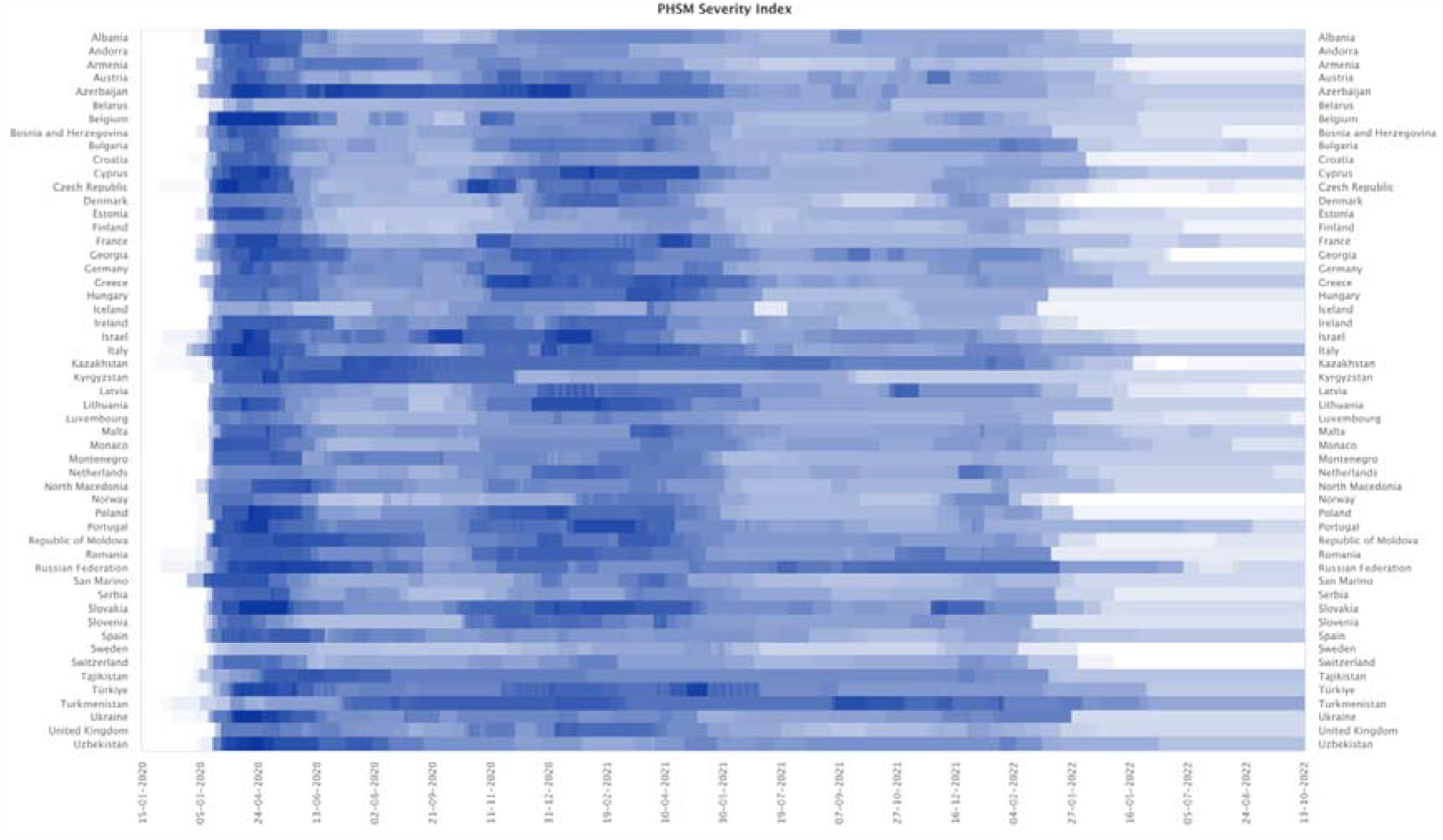
Comparison of countries’ response timing and severity – heatmap of daily PHSM Severity Index.

By quantifying qualitative PHSM data, the SI enables the analysis of the relation between PHSM severity and other types of information such as epidemiological data. PHSM and local disease epidemiology influence one another, as changes in epidemiological situations lead countries to alter their PHSM, and PHSM in turn mitigate the spread of the virus to a varying degree. At the country level, the SI can generate insights into common PHSM approaches implemented by countries over time by comparing different combinations of PHSM to the epidemiological situation, as demonstrated in Fig. 2. These visualizations enable a comparison of PHSM categories within a country and the overall responses across countries *(11)*. Cross-temporal and cross-country analyses of policy responses addressing similar epidemiological situations can help uncover the nuances of individual governments’ actions for further analysis. Specifically, countries can determine if their PHSM response is more or less severe than that of other countries with similar epidemiological situations, and explore how their PHSM responses to similar epidemiological situations have changed over time. This analysis therefore enhances understanding of how PHSM strategies at country and regional levels differ and have evolved over time.

**Figure 2.**
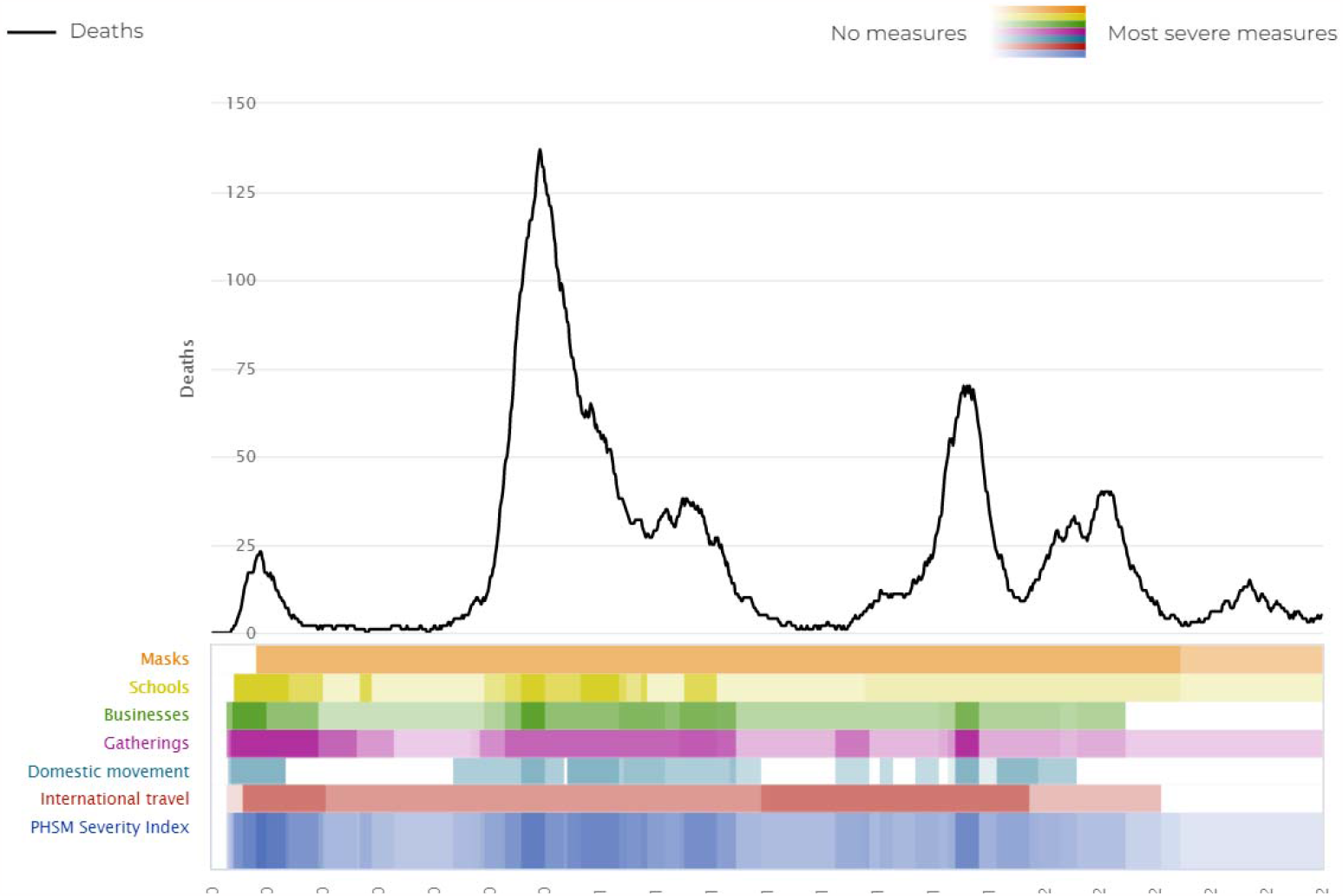
Epidemiological situation and PHSM severity (timeline) – example of country close-up: Austria. Source: PHSM in Response to COVID-19 Platform, WHO/Europe, Countries page, Austria.

Discerning the epidemiological impact of specific PHSM and combinations of measures is crucial to gain knowledge about the effectiveness of measures, and is a priority in enabling the development of evidence-informed policy *(7,18)*. WHO has launched a new multi-year initiative to measure the effectiveness and the social, health and economic impact of PHSM during health emergencies, strengthening the global evidence base on PHSM with the goal of being able to provide nuanced guidelines on the implementation of finely tuned PHSM that have a maximum effect on the mitigation of the disease, while keeping the negative effects on the society and economy at a minimum. The SI can be used to inform the research and analytical approaches addressing these questions.

To date, the SI has been used to conduct analyses of regional PHSM trends throughout the COVID-19 pandemic. Analyses have covered the topics of schools *(19,20)*, international travel *(21,22)*, masks *(23)*, businesses *(24)* and variants of concern *(25)*, which have been published by the European Observatory on Health Systems and Policies. The SI can also be used to investigate the effects of PHSM on other diseases such as tuberculosis *(26)*. In the future, the SI dataset is intended to be used by researchers for further statistical analysis of the effect of these measures on morbidity, mortality and other outcomes of interest, as has been done with other PHSM datasets *(27–29)*.

While every effort was made to ensure the accuracy and completeness of the SI through a rigorous systematic approach to data collection, categorization, coding and validation, the occurrence of some inaccuracies or incompleteness of some data is possible due to either lack of public sources, ambiguity from source wording, the translation of sources into English, inaccurate media reporting or human error. A further limitation of the SI is the fact that it only incorporates a select number of indicators of the COVID-19 response to allow for a standardized cross-country comparison and therefore does not represent an exhaustive portrayal of a country’s response. Validation exercises and the definition of coding principles helped to ensure consistency in coding severity levels and enabled the comparability of severity scores across indicators and countries. However, the measure of policy severity is inherently subjective. The overall SI score should, therefore, be understood as an approximate representation of the overall level of PHSM response active at a given time in a country, and the comparison of scores from different indicators and countries as a relative measure thereof. In addition, the index does not capture the level of enforcement and compliance with recommended and mandatory measures. Other factors that might influence adherence to PHSM, such as cultural and individual health beliefs and behaviours and vaccination uptake, may differ between countries and are also not captured by the index. When analysing behavioural change within the population, the SI should be used in combination with other indicators of population compliance that provide direct or proxy measures for social contacts, such as mobility data.

## Conclusion

Ensuring consistent, reliable and complete data during a rapidly evolving emergency is a challenge. The quality and depth of data collection for the PHSM database, which formed the foundation of the SI, was ensured by having a professional team where each member was assigned to specific countries with which they became intimately familiar and cultivated contextual expertise. In addition, direct contact with WHO country offices provided invaluable local intelligence.

The early establishment of the WHO PHSM taxonomy and glossary facilitated relevant and thorough monitoring through the categorization of measures from an early point in the pandemic. Similarly, the development of the SI methodology with accompanying coding principles was important in ensuring the consistency of application across countries by different individuals. While the structure was maintained, it was also necessary to continuously refine and adapt the methodology to best reflect the evolution of the pandemic and emerging PHSM knowledge and practices.

The COVID-19 pandemic has highlighted the importance of incorporating standardized monitoring systems and the generation of comparable data on policy responses in the emergency preparedness cycle, and should be seen as a critical component that enables the analysis of effectiveness of measures and the provision of evidence-based policy guidance. Lessons learned from the methodology and application of the SI can guide and support the development of multi-hazard PHSM monitoring and analysis frameworks for future health emergencies.

## Data Availability

All data produced in the present work are available on the WHO/EURO PHSM in Response to COVID-19 Platform.

https://phsm.euro.who.int/covid-19

## Acknowledgements

We extend our sincere appreciation to the WHO Regional Office for Europe COVID-19 Incident Management Support Team members: Caroline Brown, Calvin Cheng, Silviu Ciobanu, Ana Paula Coutinho Rehse, Ute Sylvia Enderlein, Michala Hegermann-Lindencrone, Kayla King, Alexis Luis, Lauren MacDonald, Richard Pebody, Ihor Perehinets, Jeffrey Pires, Jukka Pukkila, Celine Roman, Cristiana Salvi, Catherine Smallwood, Miranda Tran Ngoc, Paula Vasconcelos and Martin Willi Weber, who provided valuable technical expertise. We also thank our colleagues at the WHO country offices for their contribution in providing country data on public health and social measures.

